# In Vivo Mechanical Assessment of Cortical Bone Rigidity Enhances Fracture Discrimination Beyond DXA in Postmenopausal Women

**DOI:** 10.1101/2025.08.28.25334655

**Authors:** Brian C. Clark, Todd M. Manini, Janet E. Simon, Leatha A. Clark, Charalampos Lyssikatos, Stuart J. Warden

## Abstract

Dual-energy x-ray absorptiometry (DXA)-derived areal bone mineral density (BMD) remains the clinical standard for assessing osteoporosis risk, yet it fails to identify over 75% of individuals who sustain fragility fractures. Direct in vivo mechanical assessment of cortical bone strength may address this diagnostic gap by capturing structural and material properties that govern whole-bone strength but are not reflected by BMD. We conducted a multicenter case-control study with cross-sectional assessment to compare ulna flexural rigidity, a biomechanical property correlated with whole-bone strength (R² ≈ 0.99), estimated using Cortical Bone Mechanics Technology (CBMT), with DXA-derived BMD for discriminating prior fragility fractures in postmenopausal women. A total of 372 women aged 50–80 years (109 with low-trauma fractures, 263 matched controls) were enrolled across four U.S. sites. Ulna flexural rigidity was assessed by dynamic vibrational analysis; BMD was measured at the spine, hip, and 1/3 radius. Women with prior fractures had significantly lower flexural rigidity than controls (absolute: 20.0 vs. 24.8 N·m²; 21% lower; weight-normalized: 0.29 vs. 0.36 N·m²/kg; 22% lower; both P < .001). CBMT demonstrated strong discriminatory accuracy (AUC = 0.80 normalized; 0.76 absolute) versus poor DXA performance (AUC ≤ 0.63) for discriminating all fragility fractures. In multivariable models including CBMT, DXA-derived BMD, age, and BMI, CBMT remained independently associated with fracture status, whereas BMD did not. Subgroup analyses showed CBMT retained strong performance in treatment-naïve women (AUC = 0.85) and in those with non-osteoporotic BMD (AUC = 0.80). Exploratory fracture-site analyses demonstrated that ulna EI discriminated upper and lower extremity fractures, including hip, whereas DXA-derived BMD generally showed modest or nonsignificant discrimination. These findings demonstrate that in vivo mechanical assessment of cortical bone rigidity provides clinically relevant information beyond areal BMD, including women not classified high risk. Direct in vivo assessment of cortical bone rigidity may enhance fracture risk stratification and enhance osteoporosis screening.

**Lay Summary:** Most people who break a bone from a simple fall do not meet the standard definition of osteoporosis based on a bone density scan (DXA). This means many at risk are not identified or treated. Our study tested a new, noninvasive technology that directly measures how strong a bone is by assessing how much it resists bending. We found that this measure, called flexural rigidity, more accurately identified women with past fractures than DXA did, even in women whose bone density was “normal”. It also showed strong performance across different types of fractures, including hip fractures. Directly testing bone strength may help doctors better identify who needs treatment to prevent fractures.

Graphical Abstract.
Summary of The STRONGER Study design, measurement principles, and discriminatory performance of CBMT-derived ulnar flexural rigidity and DXA-derived areal bone mineral density at standard clinical sites.
**(Left)** Participant flow diagram for the STRONGER Study, a multicenter, case-control study, showing eligibility screening, adjudication, and final analytic sample (N = 372), including 109 postmenopausal women with prior fragility fractures and 263 matched controls. **(Center)** Conceptual schematics illustrating the measurement principles for Cortical Bone Mechanics Technology (CBMT) and dual-energy X-ray absorptiometry (DXA). The upper schematic depicts a simplified free-body diagram of a beam subjected to a mid-point load (*K*) over length (*L*), illustrating the Euler–Bernoulli beam theory underlying CBMT-derived calculation of ulna flexural rigidity. The lower schematic illustrates the basic principle of DXA, which measures areal bone mineral density (BMD) based on differential X-ray attenuation (recreated from Luo, Y. (2017). Bone Imaging for Osteoporosis Assessment. In: Image-Based Multilevel Biomechanical Modeling for Fall-Induced Hip Fracture. Springer, Cham. https://doi.org/10.1007/978-3-319-51671-4_3). **(Right)** Receiver operating characteristic (ROC) curves comparing the ability of CBMT-derived ulna flexural rigidity (absolute and weight-normalized) versus DXA-derived areal BMD at standard clinical sites to discriminate fracture status (all fractures). CBMT demonstrated substantially higher area under the curve (AUC) values (0.80) than any DXA site (≤0.63), highlighting its potential to capture clinically meaningful aspects of bone strength not reflected by areal BMD alone. Notably, CBMT retained strong discriminatory performance in women with non-osteoporotic BMD and more effectively identified lower extremity and hip fractures (not shown), underscoring its potential to address a critical diagnostic gap including among women not classified as high risk by conventional thresholds.

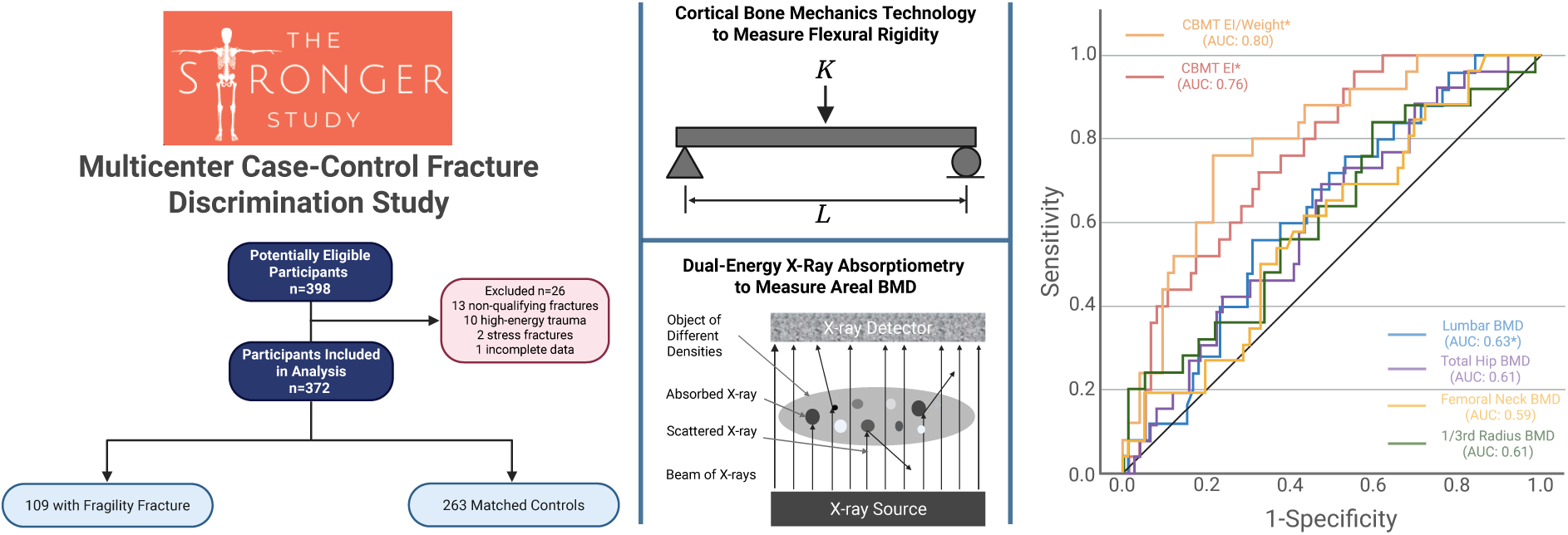

## INTRODUCTION

Osteoporosis is a skeletal disorder defined by compromised bone strength that predisposes individuals to an increased risk of fragility fractures.^(1)^ These fractures pose a major public health burden, with an estimated 2.3 million occurring each year among older adults in the United States alone, disproportionately affecting postmenopausal women.^(2,3)^ The clinical and economic impact is considerable, with related healthcare costs projected to exceed $95 billion by 2040.^(4)^

Despite these costs, the current standard for assessing fracture risk, dual-energy x-ray absorptiometry (DXA)-derived areal bone mineral density (BMD),^(5)^ has substantial limitations in its ability to identify individuals who will fracture.^(6–11)^ While areal BMD is the primary indicator for diagnosing osteoporosis, more than 75% of individuals who experience fragility fractures do not meet the diagnostic threshold for osteoporosis (T-score ≤ −2.5),^(7)^ highlighting a persistent diagnostic gap. Similarly, patients with a history of fractures remain at significantly higher risk of future fractures than those with comparable areal BMD but no prior fractures, even after adjusting for other risk factors.^(12,13)^ While bisphosphonate therapy improves bone strength and reduces fracture risk, less than 18% of the reduction in vertebral fractures can be attributed to increases in areal BMD measured by DXA.^(14,15)^ Although fracture risk calculators such as FRAX incorporate clinical risk factors to improve predictive accuracy, these tools rely heavily on areal BMD and population-level averages, and also have notable limitations in identifying high-risk individuals.^(16)^

Fundamentally, fracture resistance is governed by bone strength, a multifactorial property determined not just by bone mass but also quality, including structural (e.g., geometry, microarchitecture) and material (e.g., mineralization, collagen) properties.^(17,18)^ Areal BMD does not adequately reflect these structural and material contributors to strength, arguably contributing to its poor sensitivity for identifying individuals at high fracture risk.^(19)^ Historically, the adoption of DXA was supported by early studies suggesting strong correlations between femoral neck areal BMD and ex vivo bone strength,^(20)^ but these correlations (which had very wide confidence interval estimates) have since challenged whether areal BMD is a reasonable surrogate for mechanical strength. For example, one study recently reported that areal BMD accounted for less than 15% of the variance in key mechanical properties,^(21)^ and another showed that areal BMD explained less than 50% of femoral strength in cadaveric femurs with wide necks, with individuals having the same areal BMD showing up to a threefold difference in actual bone strength.^(22)^ Most recently, a large longitudinal study demonstrated that people with different femoral neck sizes follow distinct structural aging patterns that affect fracture risk, yet these differences are not consistently reflected in areal BMD.^(23)^ These findings reinforce that structural changes with aging vary widely within a population and that declines in areal BMD may not reliably indicate true strength loss. Taken together, these findings underscore the limitations of areal BMD in capturing clinically meaningful variation in bone strength. Improved methods are therefore needed to complement or enhance areal BMD-based assessments so that individuals at elevated fracture risk can be identified more accurately, enabling better targeting of treatment, reducing preventable fractures, and lowering related healthcare costs.

Cortical Bone Mechanics Technology (CBMT) is an emerging, noninvasive method that directly estimates mechanical whole bone strength in vivo by performing a dynamic three-point bending test of the ulna.^(6,24)^ From this, ulna flexural rigidity (EI) is derived, a biomechanical property that combines the bone’s material stiffness and structural geometry to quantify its resistance to bending. This integrated measure captures the effects of bone geometry, tissue material properties, and structural integrity, and is near-perfectly correlated with ex vivo whole-bone strength (R² ≈ 0.99).^(6,24)^ The ulna midshaft was selected as an accessible site that is almost entirely composed of cortical bone, which is especially vulnerable to age-related porosity and thinning^(25–27)^ — microstructural changes that disproportionately impact mechanical strength relative to areal BMD loss.^(28)^ CBMT-derived EI can detect weakening due to changes in structural integrity, such as collagen degradation, even when BMD remains unchanged.^(29,30)^ While CBMT does not capture all aspects of fracture resistance, such as fracture toughness or crack propagation, it provides direct, quantitative insight into the bending resistance of cortical bone — a critical contributor to whole-bone strength.

This multicenter case-control study with cross-sectional exposure measurement aimed to compare the performance of CBMT-derived ulna flexural rigidity with DXA-derived areal BMD for discriminating prior fragility fractures in postmenopausal women. We also explored two clinically relevant subgroups: women without prior osteoporosis pharmacotherapy to reduce confounding by treatment effects, and women with non-osteoporotic areal BMD (T-score > −2.5) to assess whether CBMT identifies risk missed by standard thresholds.

## METHODS

### General Overview of the Study Design and Oversight

This observational, multicenter case-control study with cross-sectional exposure measurement evaluated the ability of ulna flexural rigidity, estimated using CBMT, to discriminate fracture status in postmenopausal women. The study was conducted at four U.S. academic centers (Ohio University, Athens, OH; Indiana University–Indianapolis, Indianapolis, IN; University of Florida, Gainesville, FL; and University of Florida, Jacksonville, FL) between June 2022 and December 2023. Institutional Review Board (IRB) approval was obtained at all sites, with Ohio University serving as the IRB of record. All participants provided written informed consent before any study procedures.

The study protocol was prospectively registered at ClinicalTrials.gov (NCT05721898) and published previously.^(30)^ The design adhered to the STROBE guidelines for observational case-control studies, with independent data monitoring and oversight by the coordinating center at Ohio University. A centralized adjudication committee independently reviewed fracture documentation to confirm eligibility. Vertebral fractures were excluded because they are frequently clinically silent, often identified only incidentally on imaging, and subject to misclassification when compared with symptomatic non-vertebral fractures.^(31)^ By focusing on non-vertebral fractures, which are typically symptomatic and clinically confirmed, we strengthened the validity of our case definitions. All site personnel underwent standardized training in participant screening, consenting, and CBMT and DXA testing to ensure protocol consistency across sites.

Eligible participants were postmenopausal women aged 50–80 years, with or without a history of low-trauma fractures. Cases had to have a confirmed history of non-pathological, low-energy upper or lower limb fractures. Controls were frequency-matched on age, BMI, and race (±10%) and recruited from the same catchment areas but with no history of fragility fracture. Exclusion criteria included metabolic bone diseases other than osteoporosis, high-energy trauma fractures, and stress or insufficiency fractures. Full details of CBMT and DXA measurement protocols, including device calibration, signal quality thresholds, and operator training, are provided in the protocol publication.^(30)^

### Participants

Postmenopausal women aged 50 to 80 years were enrolled at each site. Fracture cases were defined as individuals with a history of low-trauma (fragility) fractures of the arm, leg, or pelvis sustained after age 50. Vertebral fractures were not included for the reasons noted above. Fragility fractures were operationally defined as self-reported fractures resulting from a fall from standing height or less (i.e., <6 inches). Fractures due to high-energy trauma (e.g., motor vehicle accidents, sports, etc.), stress fractures, and insufficiency fractures were excluded. Controls were required to have no history of arm or leg fracture after age 40 and were frequency-matched to cases on age, BMI, and race at a 2:1 control-to-case ratio. Complete details on the inclusion and exclusion criteria for study participants in both target populations (fragility fracture cases and controls) are provided in Supplemental Table 1. Fracture characteristics among case participants is described in Supplemental Table 2. All reported fractures were adjudicated by an independent, blinded committee using prespecified criteria and available self-reported and medical documentation. All enrolled participants underwent both CBMT and DXA testing at the study visit.

### CBMT Assessment

CBMT is a noninvasive, in vivo technology that estimates ulna flexural rigidity using a dynamic three-point bending test based on Euler–Bernoulli beam theory.^(32)^ This principle holds that bending a beam induces tensile and compressive stresses that generate a bending moment, a well-established method for quantifying structural stiffness in engineering and material science.^(32)^ The ulna midshaft was selected because of its high cortical content and its sensitivity to age-related thinning and porosity,^(25–27)^, which are key determinants of bone strength.^(28)^ Figure 1 illustrates the conceptual framework and cortical bone relevance for CBMT testing. It should be noted that an earlier approach, mechanical response tissue analysis (MRTA),^(33)^ used vibration-based flexural rigidity estimates but faced notable limitations with probe placement and mechanical support stability.^(24)^ Still, MRTA provided clear proof of concept, showing strong correlations with structural strength (r > 0.95),^(34,35)^ unique sensitivity to collagen degradation and fatigue undetected by BMD,^(36,37)^ and meaningful differences by age,^(38)^ disease,^(39)^ and training status,^(40)^ even when BMD did not change. CBMT addresses these limitations with robotics, multi-site sampling, and stable end conditions for a robust in vivo three-point bending test.^(24)^

**Figure 1.**
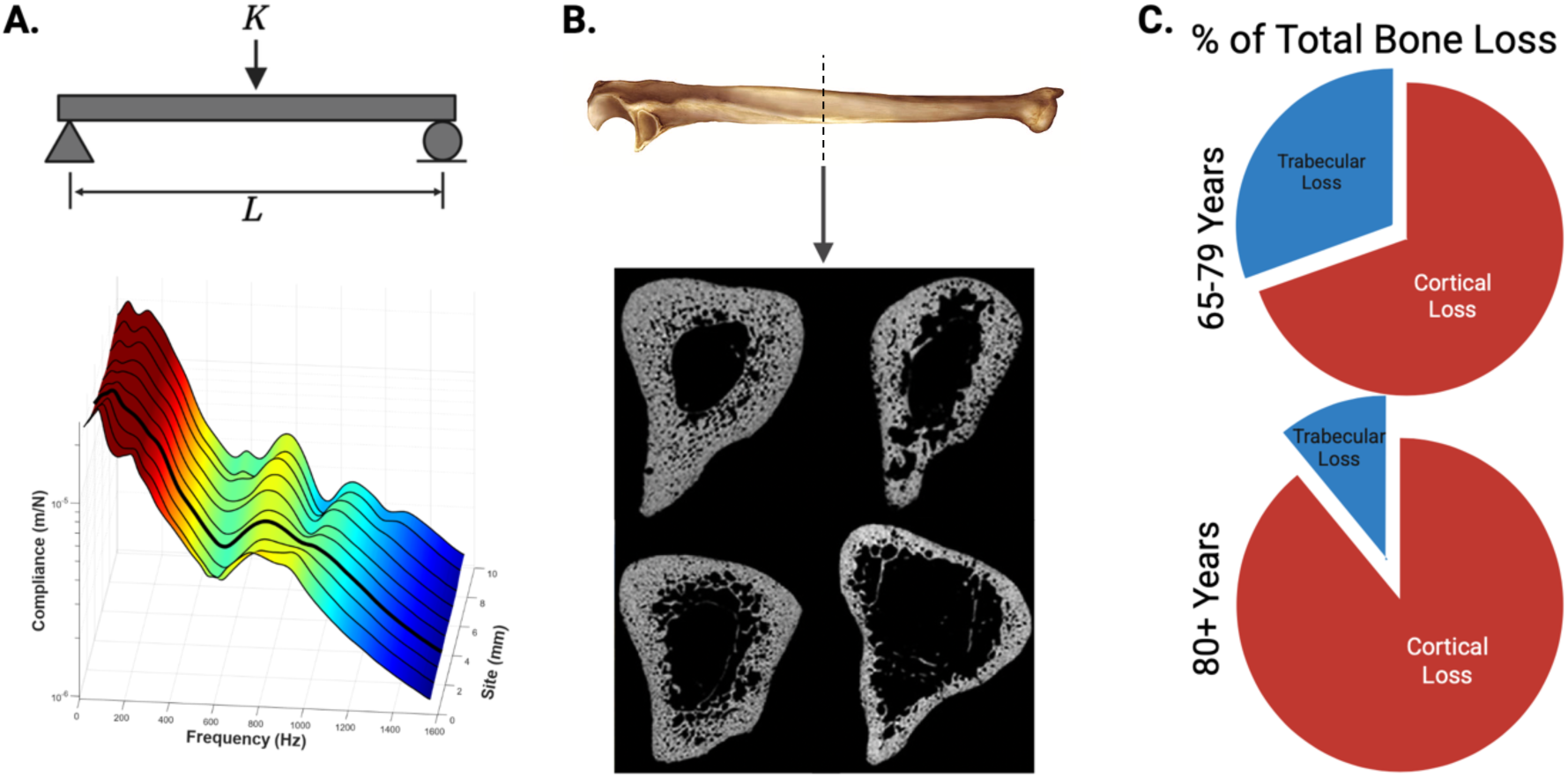
Conceptual basis and cortical bone relevance for in vivo flexural rigidity assessment. **(A)** Free-body schematic of a simply supported beam subjected to a mid-point load (*K*) over length (*L*), illustrating the fundamental principle of Euler–Bernoulli beam theory used to quantify flexural rigidity. The 3D surface plot below shows measured compliance (z-axis) across frequency (x-axis) and anatomical site index (y-axis). Data were collected from 10 adjacent sites spaced 1 mm apart to account for local variability in bone geometry and surrounding soft tissues. The color gradient depicts the magnitude of compliance, with warmer colors indicating higher values. The dark line highlights a representative high-quality frequency response function (FRF) with distinct resonance peaks, demonstrating how multi-site sampling helps capture robust bending modes despite anatomical variation. **(B)** Representative ulna bone image (Wikimedia Commons; public domain, no reprint permissions required) and cross-sectional midshaft μCT scans from four different post-mortem specimens (adapted from Bowman et al., Bone, 2019). These scans illustrate that the ulna mid-diaphysis is composed almost entirely of dense cortical bone, enclosing only a narrow central medullary canal. Although geometry and porosity naturally vary among individuals, this region consistently remains cortical-dominant. This structural consistency makes the ulna midshaft an ideal location for CBMT’s in vivo flexural rigidity assessment, as cortical bone contributes disproportionately to whole-bone strength and is particularly susceptible to age-related deterioration. **(C)** Estimated proportion of total bone loss attributed to cortical and trabecular bone with aging. These pie charts illustrate that after approximately age 65, most bone loss is cortical, which aligns with increased incidence of fragility fractures [Zebaze et al., *Lancet*, 2010]. Recreated from published data (Zebaze et al., *Lancet*, 2010).

Assessments were performed on the non-dominant arm to reduce localized loading effects and better reflect systemic skeletal health. Before testing, the ulna length (L) was measured with precision electronic calipers (0.01 mm resolution), and the mid-point was marked. Participants lay supine on the CBMT table with the forearm stabilized using wrist stabilizers and an elbow positioner to isolate the ulna from shoulder and wrist motion. Proper positioning was verified by trained operators as described in the published protocol.^(30)^

The CBMT end effector consists of a mechanical shaker, impedance head, and ceramic concave contact probe. It applies a static preload (12 N) followed by a superimposed band-limited (20–1600 Hz) vibration signal. Force and acceleration were sampled at 16 kHz and processed using proprietary MATLAB-based software, which produced an FRF that characterizes how the bone-skin system responds to applied force.

To address local variability in anatomy and minimize the effects of probe misalignment, the protocol sampled 10 discrete sites along the ulna’s diaphysis, spaced at 1 mm intervals to span a total length of 9 mm. Each site was tested under three vibration amplification conditions. This spatial sampling approach is rooted in classical beam theory and recognizes that long bones, like engineered beams, exhibit non-uniform material and geometric properties along their length. Local sampling mitigates the risk of collecting distorted signals caused by torsional or off-axis bending modes and increases the likelihood of capturing a robust FRF with clear modal peaks.

Flexural rigidity (EI) was calculated in N·m² as the product of Young’s modulus (E) and the second moment of area (I). K_B_, the dynamic bending stiffness, was derived from the FRF using two complementary fitting approaches: (1) Global fitting, which applied a two-degree-of-freedom dynamic beam model across the entire frequency range, using a weighted average of the top three sites based on the correlation to a simplified damped resonance model fit of the bone peak; and (2) Localized fitting, which employed polynomial peak extraction to isolate resonant regions for focused modeling, selecting the single site with the lowest relative model fit error. Final EI values were computed as the mean of both estimates; an approach supported by unpublished cadaver validation data demonstrating improved robustness and reduce outlier sensitivity. The final EI estimate was calculated using the standard closed-form solution for a simply supported beam under a central point load EI=(K_B_×L^3^)/48,^(41)^ and then adjusted by applying a 30% reduction to account for previously observed systematic offsets between in vivo (whole arm) measures and destructively tested cadaveric ulnae (M_peak_).^(29)^

Setup and testing took approximately 12–15 minutes. All CBMT devices were calibrated using phantom rods prior to study initiation. Operators underwent standardized training and certification. Test data were reviewed in a blinded fashion by a central coordinating center. No CBMT data were excluded. Additional technical details, probe specifications, and signal processing workflows, are provided in prior work.^(30,42)^

### DXA Assessment

Areal BMD was measured by DXA at standard clinical sites: lumbar spine (L1–L4), non-dominant hip (total hip and femoral neck), and non-dominant forearm (1/3 radius), following International Society for Clinical Densitometry guidelines.^(43)^ Sites containing surgical hardware were excluded. All scans were performed by certified technicians using standardized participant positioning and manufacturer-recommended acquisition and analysis protocols.

To ensure site-to-site comparability, all DXA densitometers were cross-calibrated at study outset using a shared Hangartner phantom (BMIL QA/QC phantom; Biomedical Imaging Laboratory, Dayton, OH).^(44)^ Each machine underwent ten repeated phantom scans, with BMD quantified in a 3.65 cm² region of interest in the center of each hydroxyapatite block. Linear regression harmonization equations were derived to align BMD measurements across the four study sites.

Daily quality assurance (QA) checks were performed per manufacturer recommendations, and weekly site-specific phantom scans (Hangartner phantom) were conducted to verify scanner stability. BMD values were required to remain within ±1.5% of the phantom baseline. If a phantom value fell outside this tolerance range, a rescan was performed; if still out of range, the machine was serviced and recalibrated according to the cross-calibration procedure above.

All DXA operators were trained using a standardized operating procedures manual and certified before study initiation. Consistent with ISCD best practices,^(45)^ all study scans were reviewed by the coordinating center in a blinded manner to ensure adherence to positioning and analysis protocols. Feedback and reanalysis requests were provided to sites as needed. When required, scans were repeated to ensure quality; if rescans were not feasible, data were excluded from final analyses.

### Blinding and Independence of Analyses

Across all study sites, DXA and CBMT analyses were conducted independently and under blinded conditions. DXA scans were analyzed by trained personnel without access to CBMT results, while CBMT outcomes were generated through a fully automated process and managed by personnel who were blinded to both DXA results and fracture status. At one site (Indiana University), the same operators performed both CBMT and DXA procedures; however, this did not affect analytic independence, as all analyses remained blinded.

### Additional Measures

Physical function was assessed using handgrip strength, usual gait speed, the Timed Up and Go test, and the Four-Square Step Test, following procedures described in the published study protocol.^(30)^ No adverse events or serious adverse events related to study procedures occurred during the study period.

### Sample Size and Power

The a priori sample size and power calculation were described in the published STRONGER protocol.^(30)^ Briefly, the primary objective was to test whether the AUC of CBMT-derived ulnar flexural rigidity exceeded 0.70 (null = 0.60) with a 2:1 ratio of controls to cases. Power analyses indicated that 190 controls and 96 cases would provide 80% power at α = 0.05. Our final sample exceeded these thresholds, ensuring adequate power for the primary analyses, though we acknowledge that certain subgroup comparisons remained less well powered.

### Statistical Analysis

Descriptive statistics were calculated for all variables. Between-group differences (fracture vs. control) were assessed using independent-samples t tests and Cohen’s d effect sizes for continuous variables, and chi-square tests and the Phi coefficient for categorical variables. Pearson correlation coefficients were used to evaluate associations between flexural rigidity and areal BMD. Normality of continuous variables was assessed by visual inspection of histograms and Q–Q plots and no substantial deviations were observed.

The primary analysis evaluated the ability of CBMT-derived ulna flexural rigidity to discriminate fracture status. The dataset was randomly partitioned into a 70% training set (N = 267) and a 30% test set (N = 105). Descriptive characteristics of the training and test subgroups were reviewed and showed no statistically significant differences in age, BMI, EI, or areal BMD measures (all p > .15), supporting the validity of the random split. Logistic regression models were developed in the training set, and predicted probabilities were applied to the test set to generate receiver operating characteristic (ROC) curves and calculate area under the curve (AUC) values. Odds ratios (ORs) and 95% confidence intervals (CIs) were estimated using 10,000 bootstrap iterations with bias-corrected and accelerated (BCa) CIs. Bootstrapping was conducted under the assumption that the sample approximated the population distribution. Optimal classification thresholds were determined using the Youden index, and sensitivity, specificity, positive predictive value (PPV), and negative predictive value (NPV) were reported.

For comparison with current clinical practice, we also classified participants using WHO osteoporosis criteria (T-score ≤ –2.5 at the lumbar spine, total hip, or femoral neck),^(46)^ and summarized sensitivity and specificity for fracture discrimination. Exploratory CBMT thresholds were derived from ROC analyses (Youden index) and are reported.

Multivariable logistic regression models were used to evaluate the discriminatory performance of CBMT-derived flexural rigidity. One model included age and BMI as covariates, and additional models paired CBMT with areal BMD at individual skeletal sites (lumbar spine, total hip, femoral neck, and 1/3 radius). Variables included in the multivariable model were selected a priori based on clinical relevance and study objectives, specifically age and BMI as established risk factors, and areal BMD given its direct relevance to the study hypothesis. Each model was assessed independently to determine whether the addition of clinical or imaging covariates improved fracture discrimination beyond CBMT alone. Linearity of continuous predictors in the logit was assessed using the Box-Tidwell procedure, and multicollinearity was evaluated using variance inflation factors (VIFs). All observations were assumed to be independent. Univariable logistic regression models were also performed to evaluate the discriminatory performance of age and BMI alone).

Flexural rigidity was examined both in absolute terms and normalized to body weight (N·m²/kg). The rationale for normalization to body weight was based on the biomechanical principle that fracture occurs when external loads exceed bone strength.^(47)^ Body weight is a primary determinant of habitual skeletal loading and of the forces experienced during falls, and thus provides a clinically meaningful reference for contextualizing bone strength.^(48)^ Because the CBMT calculation of flexural rigidity already incorporates ulna length, weight normalization conceptually complements rather than duplicates size adjustment.

Two prespecified subgroup analyses were performed: (1) excluding participants with any prior use of osteoporosis pharmacotherapy, and (2) restricting the sample to participants with non-osteoporotic areal BMD, defined as T-scores > –2.5 at either the lumbar spine, total hip, or femoral neck, consistent with WHO diagnostic thresholds.^(46)^ Participant characteristics for these subgroups are provided in Supplemental Tables 3-5, respectively.

We also conducted subgroup analyses stratified by fracture site (upper extremity, lower extremity, and hip). Due to the smaller sample sizes in these subgroups, AUC values were calculated using the full dataset rather than the 70/30 training/test split applied in the primary analyses. Because of limited power, these analyses were restricted to AUC estimation rather than full multivariable modeling or classification metrics. They provide additional insight into site-specific discriminatory performance of ulna EI relative to areal BMD, though results should be interpreted cautiously.

Missing data were minimal and handled via listwise deletion. All statistical tests were two-sided, with statistical significance defined as P < .05. Analyses were performed using SPSS version 29.0.2.0 (IBM Corp, Armonk, NY).

## RESULTS

### Participant Characteristics

Of 398 enrolled participants, 26 were excluded following adjudication (13 due to non-qualifying fracture sites, 2 due to stress or insufficiency fractures, 10 due to high-energy trauma, and 1 due to incomplete documentation), resulting in 372 participants (93% of enrolled): 109 fracture cases and 263 controls (Figure 2). Although the study was designed to recruit controls at a 2:1 ratio to cases, the final analytic sample deviated slightly from this ratio due to these adjudication-based exclusions. The mean (SD) age was 66.8 (6.2) years, and BMI was 25.9 (4.0) kg/m², with no significant differences between groups (Table 1). Racial composition was predominantly White (>90% in both groups). Fracture cases had slightly lower grip strength (P = .03) and slower Four-Square Step Test times (P = .02). Details on fracture characteristics and prior osteoporosis pharmacotherapy exposure among study participants are provided in Supplemental Tables 2 and 3.

**Figure 2.**
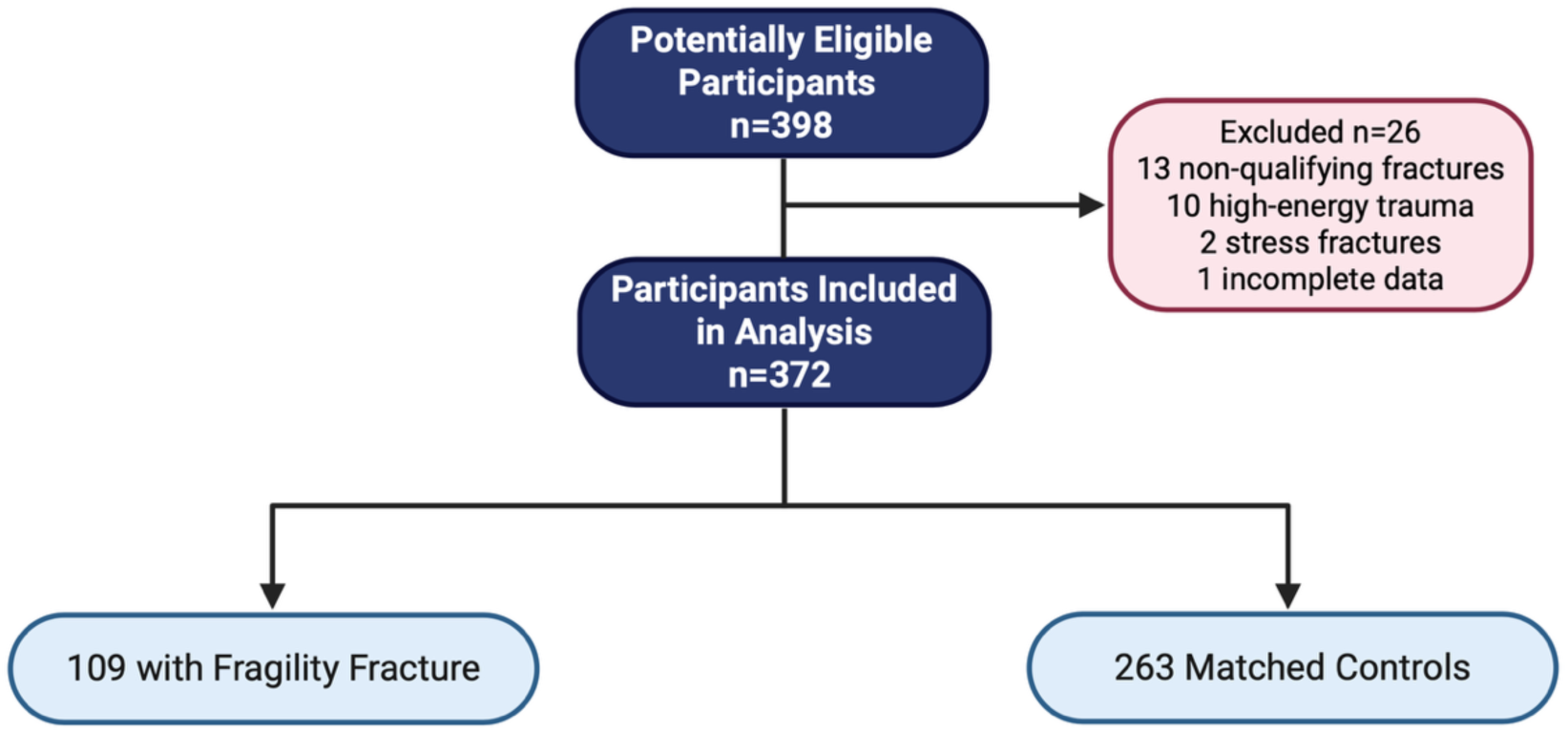
STARD flow diagram of participant inclusion. A total of 398 women were assessed for eligibility. Twenty-six were excluded due to non-qualifying fracture sites (n = 13), high-energy trauma (n = 10), stress fractures (n = 2), or incomplete documentation (n = 1). The final analytic sample included 372 postmenopausal women, with 109 fracture cases and 263 matched controls.

**Table 1.**
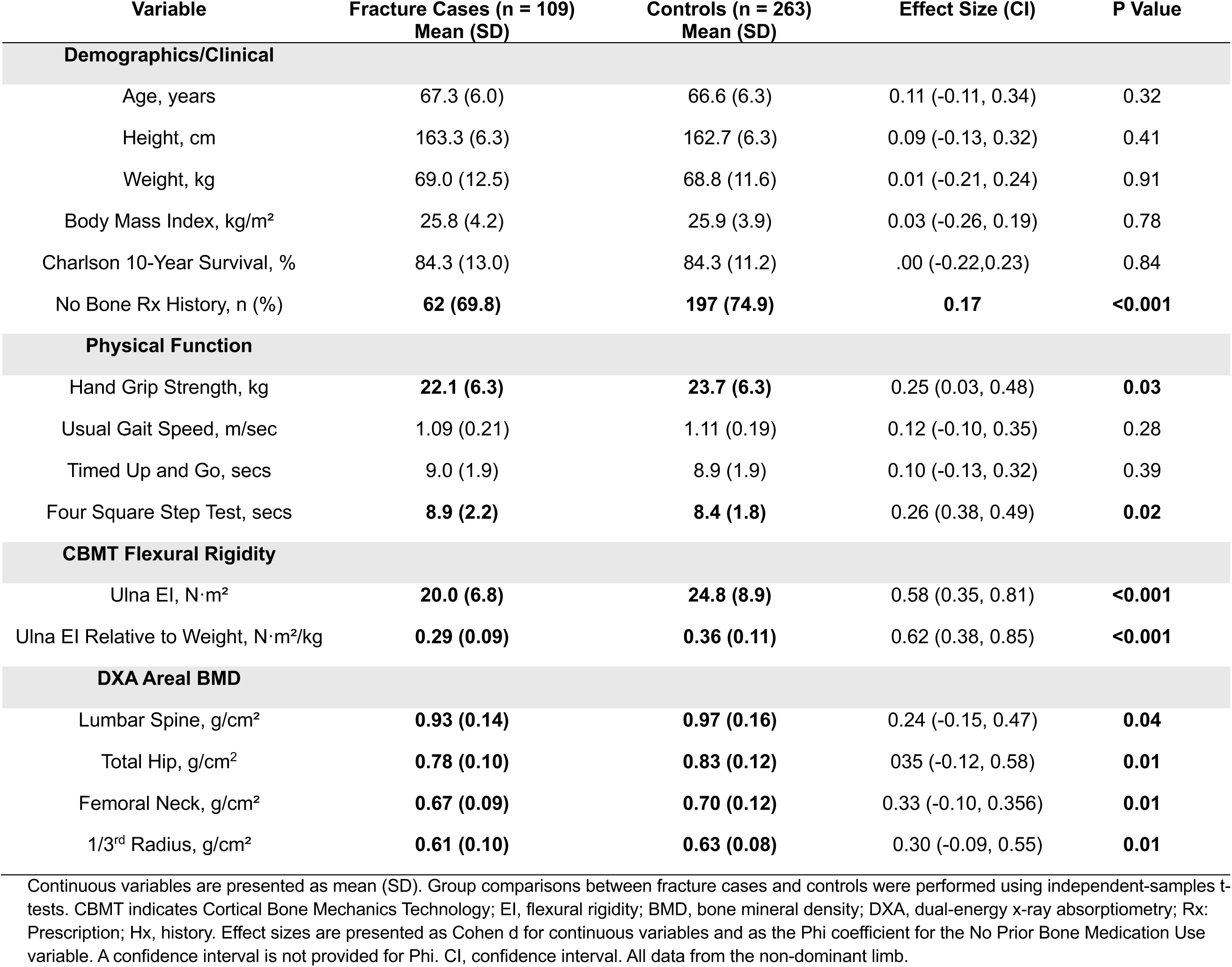
Participant Characteristics by Fracture Status in the Full Sample.

| Variable | Fracture Cases (n = 109)<br>Mean (SD) | Controls (n = 263)<br>Mean (SD) | Effect Size (CI) | P Value |
| --- | --- | --- | --- | --- |
| **Demographics/Clinical** |  |  |  |  |
| Age, years | 67.3 (6.0) | 66.6 (6.3) | 0.11 (-0.11, 0.34) | 0.32 |
| Height, cm | 163.3 (6.3) | 162.7 (6.3) | 0.09 (-0.13, 0.32) | 0.41 |
| Weight, kg | 69.0 (12.5) | 68.8 (11.6) | 0.01 (-0.21, 0.24) | 0.91 |
| Body Mass Index, kg/m<sup>2</sup> | 25.8 (4.2) | 25.9 (3.9) | 0.03 (-0.26, 0.19) | 0.78 |
| Charlson 10-Year Survival, % | 84.3 (13.0) | 84.3 (11.2) | .00 (-0.22, 0.23) | 0.84 |
| No Bone Rx History, n (%) | **62 (69.8)** | **197 (74.9)** | **0.17** | **<0.001** |
| **Physical Function** |  |  |  |  |
| Hand Grip Strength, kg | **22.1 (6.3)** | **23.7 (6.3)** | 0.25 (0.03, 0.48) | **0.03** |
| Usual Gait Speed, m/sec | 1.09 (0.21) | 1.11 (0.19) | 0.12 (-0.10, 0.35) | 0.28 |
| Timed Up and Go, secs | 9.0 (1.9) | 8.9 (1.9) | 0.10 (-0.13, 0.32) | 0.39 |
| Four Square Step Test, secs | **8.9 (2.2)** | **8.4 (1.8)** | 0.26 (0.38, 0.49) | **0.02** |
| **CBMT Flexural Rigidity** |  |  |  |  |
| Ulna EI, N·m<sup>2</sup> | **20.0 (6.8)** | **24.8 (8.9)** | 0.58 (0.35, 0.81) | **<0.001** |
| Ulna EI Relative to Weight, N·m<sup>2</sup>/kg | **0.29 (0.09)** | **0.36 (0.11)** | 0.62 (0.38, 0.85) | **<0.001** |
| **DXA Areal BMD** |  |  |  |  |
| Lumbar Spine, g/cm<sup>2</sup> | **0.93 (0.14)** | **0.97 (0.16)** | 0.24 (-0.15, 0.47) | **0.04** |
| Total Hip, g/cm<sup>2</sup> | **0.78 (0.10)** | **0.83 (0.12)** | 0.35 (-0.12, 0.58) | **0.01** |
| Femoral Neck, g/cm<sup>2</sup> | **0.67 (0.09)** | **0.70 (0.12)** | 0.33 (-0.10, 0.356) | **0.01** |
| 1/3<sup>rd</sup> Radius, g/cm<sup>2</sup> | **0.61 (0.10)** | **0.63 (0.08)** | 0.30 (-0.09, 0.55) | **0.01** |

Continuous variables are presented as mean (SD). Group comparisons between fracture cases and controls were performed using independent-samples t-tests. CBMT indicates Cortical Bone Mechanics Technology; EI, flexural rigidity; BMD, bone mineral density; DXA, dual-energy x-ray absorptiometry; Rx: Prescription; Hx, history. Effect sizes are presented as Cohen d for continuous variables and as the Phi coefficient for the No Prior Bone Medication Use variable. A confidence interval is not provided for Phi. CI, confidence interval. All data from the non-dominant limb.

### Discriminatory Accuracy of CBMT for All Fractures

CBMT-derived ulna flexural rigidity was significantly lower in fracture cases than in controls (20.0 [6.8] vs 24.8 [8.9] N·m²; P < .001) (Table 1, Figure 3). This difference persisted after normalization to body weight (0.29 [0.09] vs 0.36 [0.11] N·m²/kg; P < .001) (Table 1, Figure 3). In the full sample, the AUC for absolute flexural rigidity was 0.76 (95% CI, 0.66–0.86) and improved to 0.80 (95% CI, 0.70–0.89) when normalized to body weight (Table 2; Figure 4A). The optimal threshold for weight-normalized EI (0.27 N·m²/kg) yielded 76.0% sensitivity and 78.4% specificity (Table 2). All reported discriminatory metrics reflect performance in the independent test set. As shown in Supplemental Table 6, age and BMI alone were not significant discriminators of fracture status (AUCs <0.60), reinforcing the limited utility of conventional clinical risk factors and underscoring the need for more direct assessments of bone strength.

**Figure 3.**
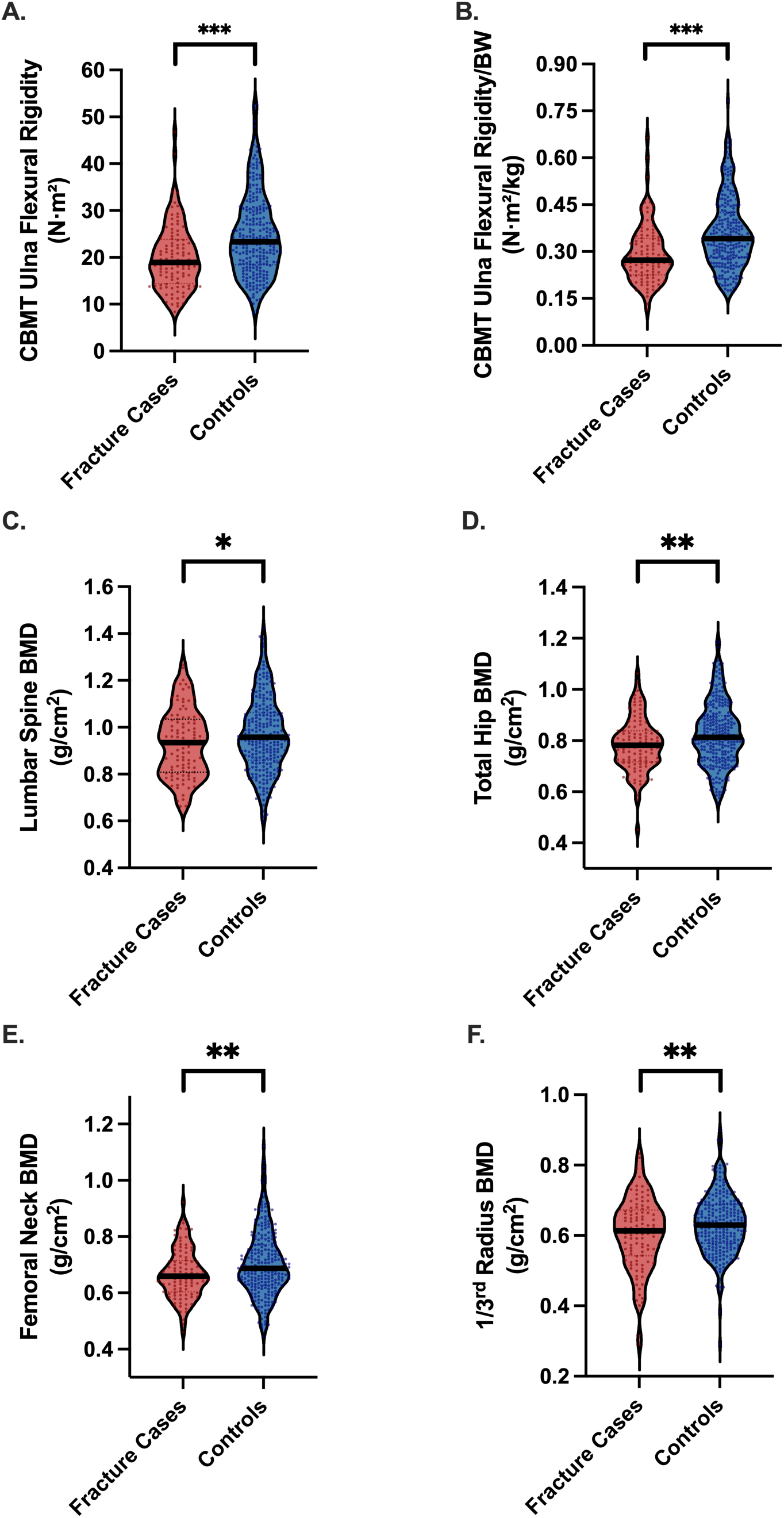
Differences in CBMT Flexural Rigidity (EI) and DXA-Derived areal Bone Mineral Density (BMD) Between Fracture Cases and Controls. The percent difference in EI between fracture cases and controls was approximately 22%, compared with only 3–6% for BMD measures. **Panels A and B** display cortical bone mechanical ulnar flexural rigidity measured using CBMT, expressed in absolute terms (A) and normalized to body weight (B). **Panels C through F** present DXA-derived areal BMD at the lumbar spine (C), total hip (D), femoral neck (E), and 1/3 radius (F). All measurements, except lumbar spine BMD, were obtained from the non-dominant limb. Asterisks indicate statistically significant group differences (*p < 0.05; ** p < 0.01; ***p < 0.001).

**Figure 4.**
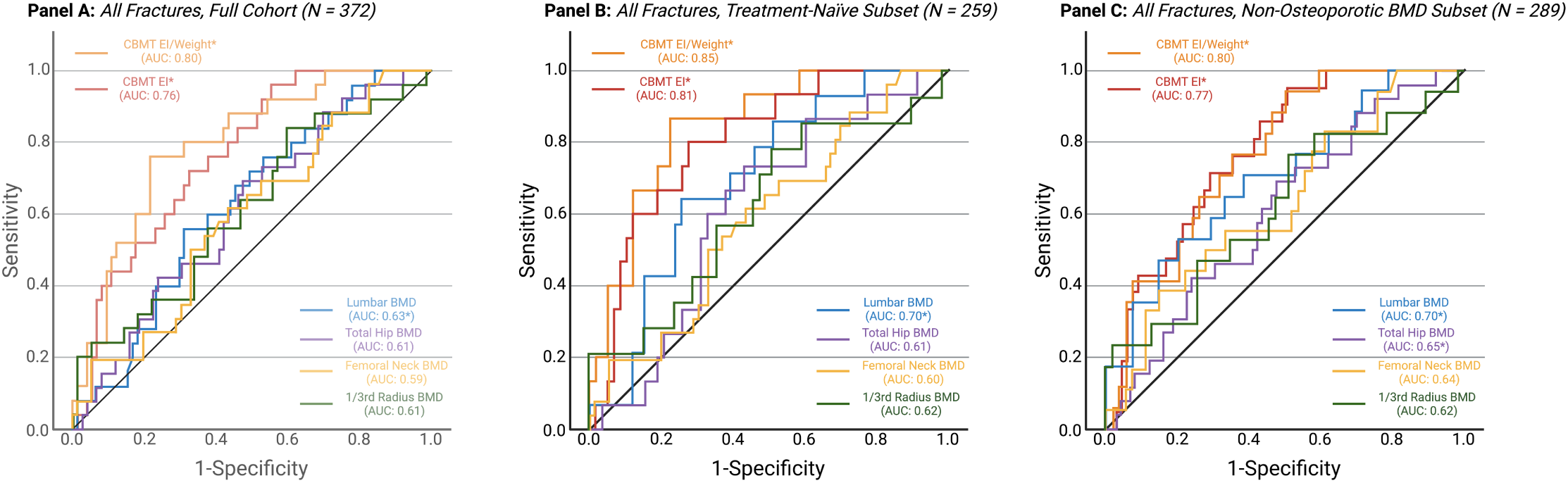
ROC Curves Comparing CBMT Flexural Rigidity and DXA-Derived Areal Bone Mineral Density (BMD) for Discriminating Fracture Status. Panel. **A** shows receiver operating characteristic (ROC) curves for the full sample (N = 372), comparing CBMT-derived ulna flexural rigidity—expressed in absolute terms and normalized to body weight—with DXA-derived areal BMD at the lumbar spine, total hip, femoral neck, and 1/3 radius. CBMT demonstrated AUCs of 0.76 (absolute) and 0.80 (normalized) (P < .001), indicating strong discriminatory performance. In contrast, AUCs for DXA sites ranged from 0.59 to 0.63, with only the lumbar spine being statistically significant in univariable models. **Panel B** presents results from the subset excluding participants with prior osteoporosis pharmacotherapy (n = 259). CBMT performance improved, with AUCs of 0.81 (absolute) and 0.85 (normalized), while areal BMD AUC’s were notably lower (0.60-0.70). **Panel C** shows ROC curves for the subset of participants with non-osteoporotic areal BMD (T-score > –2.5 at the lumbar spine, total hip, or femoral neck; n = 289). CBMT retained strong discriminatory performance (AUC = 0.77 for absolute and 0.80 for normalized), whereas DXA-derived areal BMD again showed modest discrimination (AUCs ≤ 0.70). Additional analyses stratified by fracture site demonstrated that CBMT also retained discriminatory performance for both lower extremity and hip fractures, while DXA-derived BMD continued to perform poorly (not shown; see results for details).

**Table 2.** Logistic Regression and AUC Results for CBMT and DXA in Discriminating Fracture Status Across Full and Stratified Cohorts. Threshold-based metrics (cutoff, sensitivity, specificity, PPV, NPV) are omitted for predictors with non-significant discriminatory performance (AUC P > .05).

| Logistic Regression Results |  |  |  | ROC Curve Results |  |  |  |  |  |  |  |
| --- | --- | --- | --- | --- | --- | --- | --- | --- | --- | --- | --- |
| Predictor | OR<br>Per SD | 95% CI | P-Value | AUC | 95% CI | P-Value | Cutoff | Sensitivity<br>(%) | Specificity<br>(%) | PPV<br>(%) | NPV<br>(%) |
| **Full Sample** |  |  |  |  |  |  |  |  |  |  |  |
| CBMT Absolute EI | **0.58** | **0.43, 0.81** | **< 0.001** | **0.76** | **0.66, 0.86** | **< 0.001** | **25.7 N·m<sup>2</sup>** | **96.0** | **44.6** | **36.9** | **97.1** |
| CBMT EI Relative to Weight | **0.57** | **0.42, 0.80** | **< 0.001** | **0.80** | **0.70, 0.89** | **< 0.001** | **0.27 N·m<sup>2</sup>/kg** | **76.0** | **78.4** | **54.3** | **90.9** |
| Lumbar Areal BMD | 0.85 | 0.64, 1.11 | 0.23 | **0.63** | **0.51, 0.74** | **0.04** | **0.89 g/cm<sup>2</sup>** | **56.0** | **68.8** | **37.1** | **82.6** |
| Total Areal Hip BMD | **0.72** | **0.54, 0.95** | **0.02** | 0.61 | 0.49, 0.73 | 0.07 | — | — | — | — | — |
| Femoral Neck Areal BMD | **0.69** | **0.52, 0.93** | **<0.01** | 0.59 | 0.47, 0.72 | 0.15 | — | — | — | — | — |
| 1/3<sup>rd</sup> Radius Areal BMD | 0.78 | 0.59, 1.05 | 0.10 | 0.61 | 0.49, 0.74 | 0.08 | — | — | — | — | — |
| **Treatment-Naïve Subset** |  |  |  |  |  |  |  |  |  |  |  |
| CBMT Absolute EI | **0.67** | **0.46, 0.99** | **0.04** | **0.81** | **0.69, 0.92** | **< 0.001** | **18.59 N·m<sup>2</sup>** | **80.0** | **72.4** | **42.9** | **93.8** |
| CBMT EI Relative to Weight | **0.55** | **0.36, 0.84** | **0.01** | **0.85** | **0.75, 0.95** | **< 0.001** | **0.27 N·m<sup>2</sup>/kg** | **86.7** | **77.6** | **50.0** | **96.0** |
| Lumbar Areal BMD | 0.72 | 0.49, 1.06 | 0.08 | **0.70** | **0.57, 0.84** | **0.01** | **0.88 g/cm<sup>2</sup>** | **64.3** | **74.6** | **38.4** | **89.5** |
| Total Hip Areal BMD | **0.60** | **0.41, 0.88** | **0.01** | 0.61 | 0.47, 0.76 | 0.13 | — | — | — | — | — |
| Femoral Neck Areal BMD | **0.65** | **0.44, 0.95** | **0.02** | 0.60 | 0.45, 0.75 | 0.20 | — | — | — | — | — |
| 1/3<sup>rd</sup> Radius Areal BMD | 0.79 | 0.52, 1.19 | 0.23 | 0.62 | 0.45, 0.79 | 0.16 | — | — | — | — | — |
| **Non-Osteoporotic BMD Subset** |  |  |  |  |  |  |  |  |  |  |  |
| CBMT Absolute EI | **0.57** | **0.38, 0.84** | **<0.01** | **0.77** | **0.65, 0.88** | **< 0.001** | **25.71 N·m<sup>2</sup>** | **94.1** | **50.0** | **37.3** | **96.4** |
| CBMT EI Relative to Weight | **0.55** | **0.36, 0.83** | **<0.01** | **0.80** | **0.69, 0.92** | **< 0.001** | **0.39 N·m<sup>2</sup>/kg** | **70.6** | **83.3** | **57.2** | **90.0** |
| Lumbar Areal BMD | 0.87 | 0.63, 1.20 | 0.38 | **0.70** | **0.56, 0.85** | **<0.01** | **0.92 g/cm<sup>2</sup>** | **52.9** | **80.0** | **45.5** | **84.3** |
| Total Hip Areal BMD | **0.70** | **0.51, 0.96** | **0.02** | **0.65** | **0.51, 0.79** | **0.04** | **0.81 g/cm<sup>2</sup>** | **55.6** | **74.1** | **40.4** | **84.1** |
| Femoral Neck Areal BMD | **0.57** | **0.38, 0.87** | **<0.01** | **0.64** | **0.49, 0.78** | **0.07** | — | — | — | — | — |
| 1/3<sup>rd</sup> Radius Areal BMD | 0.90 | 0.64, 1.28 | 0.56 | 0.62 | 0.46, 0.78 | 0.14 | — | — | — | — | — |

Odds ratios (ORs) are expressed per 1–standard deviation (SD) decrease in the predictor. ROC = Receiver operating characteristic curve. AUC = area under the ROC curve. CBMT = Cortical Bone Mechanics Technology; EI = Flexural rigidity; BMD = bone mineral density. The cutoff value was derived from the Youden Index. All data from the non-dominant limb.

### DXA Discrimination and Comparative Performance for All Fractures

Areal BMD values at all sites were lower in all fracture cases, with statistical significance observed only at the lumbar spine (Table 1, Figure 3). ROC analysis demonstrated modest discrimination: AUCs for lumbar spine, total hip, femoral neck, and 1/3 radius were 0.63, 0.61, 0.59, and 0.61, respectively (Table 2, Figure 4A).

In multivariable models that included absolute CBMT-derived EI and adjusted for age and BMI, CBMT-derived EI remained significantly associated with fracture status (OR = 0.59; 95% CI, 0.41–0.84; P < .01), while age and BMI were not significant predictors (Table 3; Supplemental Table 7 presents ORs and ROC results for flexural rigidity normalized to body weight CBMT EI values). The AUC for the CBMT + age + BMI model was 0.76 (95% CI, 0.66– 0.86), slightly less than CBMT alone (Table 3), indicating that conventional risk factors did not add predictive value beyond CBMT.

**Table 3.** Multivariable Logistic Regression Models Predicting Fracture Status Using Absolute CBMT-Derived Flexural Rigidity with Adjustment for Age, BMI, and DXA-Derived areal BMD in the Full Sample.

| Model | Predictors | Logistic Regression Results |  |  | ROC Curve Results |  |  |
| --- | --- | --- | --- | --- | --- | --- | --- |
|  |  | OR Per SD | 95% CI | P-Value | AUC | 95% CI | P-Value |
| **Absolute CBMT EI Only (Univariable)** |  |  |  |  | **0.76** | **0.66–0.86** | **< 0.001** |
|  | Absolute CBMT EI | **0.58** | **0.43, 0.81** | **< 0.001** |  |  |  |
| **Absolute CBMT EI + Age and BMI** |  |  |  |  | **0.76** | **0.66-0.86** | **<0.001** |
|  | Absolute CBMT EI | **0.59** | **0.41, 0.84** | **<0.01** |  |  |  |
|  | Age | 1.06 | 0.79, 1.41 | 0.71 |  |  |  |
|  | BMI | 1.01 | 0.73, 1.41 | 0.94 |  |  |  |
| **Absolute CBMT EI + Lumbar Areal BMD** |  |  |  |  | **0.75** | **0.65, 0.85** | **< 0.001** |
|  | Absolute CBMT EI | **0.60** | **0.41, 0.83** | **< 0.01** |  |  |  |
|  | Lumbar Areal BMD | 0.90 | 0.78, 1.04 | 0.49 |  |  |  |
| **Absolute CBMT EI + Total Hip Areal BMD** |  |  |  |  | **0.74** | **0.64, 0.84** | **< 0.001** |
|  | Absolute CBMT EI | **0.66** | **0.48, 0.91** | **< 0.01** |  |  |  |
|  | Total Hip Areal BMD | 0.78 | 0.58, 1.05 | 0.11 |  |  |  |
| **Absolute CBMT EI + Femoral Neck Areal BMD** |  |  |  |  | **0.73** | **0.63, 0.83** | **< 0.001** |
|  | Absolute CBMT EI | **0.67** | **0.48, 0.94** | **0.02** |  |  |  |
|  | Femoral Neck Areal BMD | 0.79 | 0.58, 1.08 | 0.13 |  |  |  |
| **Absolute CBMT EI + 1/3<sup>rd</sup> Radius Areal BMD** |  |  |  |  | **0.74** | **0.654 0.84** | **< 0.001** |
|  | Absolute CBMT EI | **0.63** | **0.45, 0.88** | **< 0.01** |  |  |  |
|  | 1/3<sup>rd</sup> Radius Areal BMD | 1.87 | 0.62, 1.23 | 0.42 |  |  |  |

Odds ratios (ORs) are expressed per 1–standard deviation (SD) decrease in the predictor. ROC = Receiver operating characteristic curve. AUC = area under the ROC curve. CBMT = Cortical Bone Mechanics Technology; EI = Flexural rigidity; BMD = bone mineral density. All data from the non-dominant limb.

When absolute CBM-derived EI was modeled alongside areal BMD from individual skeletal sites, CBMT-derived EI remained independently associated with fracture status (OR range = 0.60–0.67; all P < .02), while BMD was not (all P-values not significant) (Table 3). The addition of areal BMD did not meaningfully improve model discrimination (AUC range: 0.73– 0.75). Similar findings were observed for the multivariable models of flexural rigidity normalized to body weight (Supplemental Table 7). CBMT EI and areal BMD were only weakly correlated (r = 0.22–0.33), suggesting that the two modalities capture distinct skeletal attributes (Figure 5).

**Figure 5.**
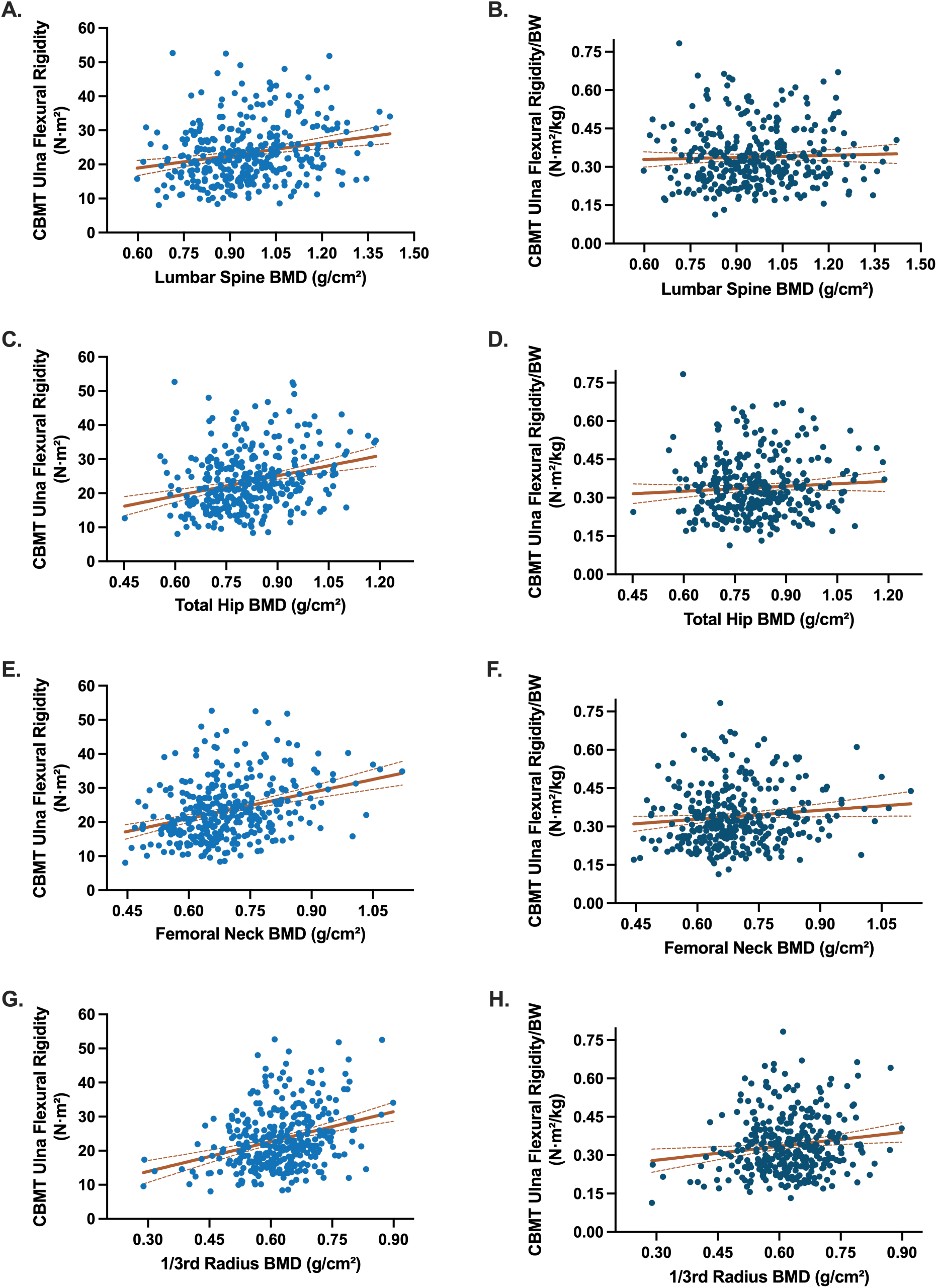
Scatterplots illustrating the relationships between CBMT-derived ulna flexural rigidity and DXA-derived areal bone mineral density (BMD) across standard clinical sites. Panels on the left show absolute flexural rigidity (N·m²; blue) plotted against areal BMD at the lumbar spine (A), total hip (C), femoral neck (E), and 1/3 radius (G). Panels on the right show weight-normalized flexural rigidity (N·m²/kg; dark blue) plotted against the same areal BMD sites: lumbar spine (B), total hip (D), femoral neck (F), and 1/3 radius (H). Trend lines indicate fitted linear regressions; Pearson correlation coefficients (r) were: Panel A (lumbar spine, absolute EI) r = 0.22, P < .001; Panel B (lumbar spine, normalized EI) r = 0.04, P = .48; Panel C (total hip, absolute EI) r = 0.28, P < .001; Panel D (total hip, normalized EI) r = 0.07, P = .20; Panel E (femoral neck, absolute EI) r = 0.33, P < .001; Panel F (femoral neck, normalized EI) r = 0.11, P = .037; Panel G (1/3 radius, absolute EI) r = 0.31, P < .001; and Panel H (1/3 radius, normalized EI) r = 0.15, P = .007.

Using WHO diagnostic criteria for clinical osteoporosis (T-score ≤ –2.5 at the lumbar spine, total hip, or femoral neck)^(46)^, only 25.7% of fracture cases were classified as osteoporotic, meaning that 74.3% were not captured by standard DXA thresholds; specificity was 82.8%. By contrast, exploratory CBMT cutoffs derived from ROC analysis (Table 2) demonstrated substantially greater sensitivity, particularly for absolute EI, though these cutpoints require prospective validation.

### Subgroup Analyses: Pharmacotherapy and Non-Osteoporotic BMD

In the treatment-naïve subgroup (n = 259; with no significant differences between groups for age or BMI as shown in Supplementary Table 3), CBMT showed improved discriminatory accuracy (AUC = 0.81 for absolute EI; 0.85 for weight-normalized EI) (Table 2, Figure 4B). Odds ratios per SD remained significant in univariable models (Table 2). Areal BMD remained non-significant across all sites with the exception of the lumbar spine (Table 2).

In the subgroup with non-osteoporotic areal BMD (n = 298; with no significant differences between groups for age or BMI as shown in Supplementary Table 5), CBMT continued to discriminate fracture status (AUC = 0.77–0.80) with substantially higher sensitivity (94% vs.53-56%), while areal BMD at the lower spine (AUC=0.70) and total hip (AUC=0.65) reached significance but remained less predictive (Table 2, Figure 4C). The latter subgroup finding demonstrates that CBMT remains robust even in women typically considered low-risk by standard criteria (i.e., those whose areal BMD values are “normal” or osteopenic).

### Subgroup Analyses: Fracture Site

When stratified by fracture site, CBMT-derived EI continued to outperform DXA-derived areal BMD. For upper extremity fractures (68 cases, 258 controls), absolute EI discriminated fracture status with an AUC of 0.66 (95% CI, 0.59–0.73; p < .001) and weight-normalized EI with an AUC of 0.68 (95% CI, 0.61–0.75; p < .001). In contrast, areal BMD values at the femoral neck (AUC: 0.58, 95% CI: 0.51, 0.65; p=0.03), total hip (AUC: 0.60; 95% CI: 0.53, 0.67; p=0.01), lumbar spine (AUC: 0.57; 95% CI: 0.50, 0.65; p=0.07), and 1/3^rd^ radius (AUC: 0.62; 95% CI: 0.54-0.70; p=0.01) all demonstrated lower discrimination.

For lower extremity fractures (40 cases, 258 controls), absolute EI again showed superior performance (AUC = 0.67; 95% CI, 0.58–0.76; p < .001), similar to weight-normalized EI (AUC = 0.67; 95% CI, 0.58–0.76; p < .001). In contrast, areal BMD values at the femoral neck (AUC: 0.60, 95% CI: 0.51, 0.69; p=0.04), total hip (AUC: 0.57; 95% CI: 0.47, 0.67; p=0.17), lumbar spine (AUC: 0.54; 95% CI: 0.45, 0.64; p=0.36), and 1/3^rd^ radius (AUC: 0.48; 95% CI: 0.39-0.57; p=0.68) all demonstrated lower discrimination.

Finally, for hip fractures (9 cases, 258 controls), absolute EI demonstrated strong discriminatory capacity (AUC = 0.77; 95% CI, 0.64–0.90; p < .001). Weight-normalized EI performed less well (AUC = 0.65; 95% CI, 0.49–0.81; p = .072). In contrast, all areal BMD measures showed non-significant results, with the femoral neck (AUC: 0.64, 95% CI: 0.44, 0.85; p=0.17), total hip (AUC: 0.69; 95% CI: 0.48, 0.90; p=0.07), lumbar spine (AUC: 0.45; 95% CI: 0.45, 0.64; p=0.36), and 1/3^rd^ radius (AUC: 0.46; 95% CI: 0.32-0.60; p=0.57) all demonstrating non-significant discriminatory performance.

## DISCUSSION

In this multicenter case-control study, we found that mechanical estimation of cortical bone strength using CBMT EI significantly outperformed DXA-derived areal BMD in discriminating postmenopausal women with and without prior fragility fractures. CBMT remained independently associated with fracture status in all multivariable models, whereas no DXA site demonstrated significant discriminatory utility. Together, these findings build on recent evidence demonstrating the limitations of areal BMD alone and suggest that direct in vivo mechanical assessment may improve fracture risk stratification and inform more targeted intervention strategies.

Fracture resistance depends on both bone quantity and quality, including material properties and structural geometry, features not adequately captured by areal BMD.^(17,18)^ Unlike DXA, CBMT estimates a bone’s resistance to bending in vivo, integrating material and geometric characteristics into a single biomechanical property: flexural rigidity (*EI*).^(6,24)^ Previous validation studies have shown that CBMT-derived EI is highly predictive of cadaveric ulna strength (R² = 0.99) and sensitive to microstructural changes, such as collagen integrity loss, even when areal BMD remains stable.^(29,30)^ In the current study, the weak correlations between CBMT EI and areal BMD (less than 11% explained variance) reinforce that the modalities assess distinct skeletal attributes and underscore the need for complementary measures in clinical practice. An important consideration is that CBMT quantifies flexural rigidity (EI), a composite measure equal to the product of apparent elastic modulus and cross-sectional geometry. Although CBMT does not distinguish between ‘quantity’ (geometry) and ‘quality’ (material) contributions, this is arguably a clinical strength, as fracture susceptibility depends on overall bone competence rather than any single determinant.

Our results are consistent with previous work demonstrating that conventional DXA-derived areal BMD fails to identify a substantial proportion of individuals who sustain fragility fractures.^(6–11)^ For instance, in The Rotterdam Study, which followed more than 14,000 individuals for up to 20 years, over 75% of fragility fractures occurred in individuals with areal BMD values above the WHO diagnostic threshold for osteoporosis,^(7)^ highlighting a major gap in the ability to identify those truly at risk. Similarly, a meta-analysis of eight prospective cohort studies found that femoral neck BMD had high specificity but poor sensitivity, with false-negative rates exceeding 80% for fracture prediction.^(9)^ Longitudinal changes in BMD are also limited by measurement error and fail to consistently predict changes in fracture risk.^(49)^

A unique strength of CBMT lies in its focus on cortical bone: at the ulna midshaft, the bone is composed almost entirely of dense cortical tissue surrounding a narrow medullary cavity (Figure 1), which contributes disproportionately to whole-bone strength.^(25–28)^ Cortical bone is especially susceptible to age-related porosity and thinning, microstructural changes that can dramatically reduce mechanical integrity.^(25–28)^ As remodeling imbalances increase cortical porosity and reduce trabecular connectivity, these changes disproportionately weaken mechanical strength compared to the degree of BMD loss.^(28)^ This mechanistic focus may explain why CBMT demonstrated strong discriminatory capacity, with high sensitivity when expressed in absolute terms (96%) and balanced sensitivity and specificity when normalized to body weight (76% and 78%, respectively). Normalization to body weight was intended to contextualize bone strength relative to the habitual and fall-related loads the skeleton must withstand,^(47,48)^ complementing the ulna length adjustment already inherent in the CBMT calculation. Notably, when benchmarked against standard clinical DXA thresholds (T-score ≤ – 2.5), sensitivity for identifying fracture cases was only ∼26%, consistent with the well-recognized diagnostic gap of DXA. By contrast, exploratory CBMT thresholds (Table 2) identified a substantially greater proportion of fracture cases, underscoring its potential to improve clinical detection if future studies confirm optimal cutpoints.

A reasonable question is whether ulna flexural rigidity generalizes to other fracture-prone skeletal sites such as the hip, which differ in loading environment and tissue composition. Ulna EI is a direct, in vivo measure of flexural rigidity that integrates material stiffness and cross-sectional geometry to quantify resistance to bending, a well-established determinant of mechanical integrity in engineering and bone biomechanics.^(17,18)^ Our fracture-site stratified analyses showed that ulna EI retained discriminatory capacity across locations and was particularly strong for hip fractures, whereas DXA-derived BMD performed poorly across the same strata. We did not include vertebral fractures in this study given their frequent subclinical presentation and potential for misclassification, so we cannot address EI’s relevance at that site. Thus, while ulna EI should not be viewed as a direct surrogate for site-specific measures, it represents a robust system-level biomarker of cortical mechanical integrity that complements BMD by capturing aspects of bone quality and structure more directly tied to whole-bone strength.

We also conducted prespecified subgroup analyses to address potential confounding from pharmacotherapy. Osteoporosis treatment is commonly initiated after fracture and may alter bone microstructure, obscuring diagnostic differences. Notably, in the treatment-naïve subgroup, CBMT’s performance improved further (AUC = 0.85). Importantly, CBMT maintained strong discrimination among participants with non-osteoporotic BMD (T-scores > –2.5), highlighting its potential to close a critical diagnostic gap for the majority of patients who fracture despite being classified as “low risk” by BMD alone.

Compared to high-resolution peripheral quantitative computed tomography (HR-pQCT) and finite element analysis, which offer detailed microarchitectural data but are limited by cost, availability, and analytic complexity,^(50)^ CBMT provides a practical, noninvasive, and radiation-free option that directly measures a biomechanical property of structural relevance. These characteristics make CBMT well positioned for integration into clinical practice as a complementary tool alongside DXA.

Future research should explore the potential of CBMT to reclassify fracture risk prospectively in diverse populations, including men, underrepresented minorities, and individuals with comorbid conditions that affect bone health. Additionally, studies evaluating the responsiveness of CBMT-derived flexural rigidity to pharmacologic and non-pharmacologic interventions would provide critical evidence on its utility for monitoring treatment effects. Integrating CBMT into existing fracture risk algorithms (e.g., FRAX) or developing new risk calculators that combine BMD, CBMT, and clinical risk factors could further improve prediction accuracy and inform clinical decision-making.

This study has limitations. Its retrospective case-control design precludes estimation of absolute or prospective fracture risk and may be subject to recall and selection bias, despite our use of blinded adjudication based on prespecified criteria. Second, the study cohort was predominantly White and female, limiting generalizability; future validation in more racially and ethnically diverse populations is needed to determine broader applicability and clinical impact. Third, cost-effectiveness analyses are warranted to assess the practical implications of implementing CBMT in routine osteoporosis screening workflows. Finally, a further limitation relates to data quality control. Recent work has led to the development and validation of a CBMT signal quality indicator scoring system designed to reflect the fidelity of the frequency response function signal.^(42)^ At the time of this study, this metric had not yet been implemented; therefore, operators did not have a standardized protocol to determine in real time whether a given scan had adequate signal quality and should be repeated. Although this could theoretically increase measurement variability, prior analyses from the same STRONGER cohort showed that correlations between CBMT signal quality and biometric properties such as BMI, forearm circumference, and skinfold thickness were weak to very weak.^(42)^ Despite the absence of a QC repeat protocol, CBMT nevertheless demonstrated strong discriminatory performance across the cohort. Looking forward, use of the signal quality indicator as a ‘go/no-go’ tool to determine whether a scan should be repeated is expected to further enhance the clinical utility of CBMT by minimizing the potential influence of suboptimal measurements.

## CONCLUSIONS

In summary, CBMT-derived ulna flexural rigidity, a direct, in vivo mechanical estimate of cortical bone strength, was independently associated with fracture status and consistently outperformed DXA-derived areal BMD in this multicenter case-control study. CBMT retained strong discriminatory performance even among women with non-osteoporotic BMD. Importantly, fracture-site analyses showed that ulna EI discriminated fractures across skeletal locations, including lower extremity and hip hip fractures, whereas DXA-derived BMD generally performed modestly. These results support ulna EI as a system-level indicator of cortical bone mechanical integrity that complements BMD. Direct, in vivo mechanical assessment of whole bone strength may therefore enhance fracture risk stratification and help close the critical diagnostic blind spot in osteoporosis care and to guide more precise, targeted prevention and treatment strategies.

## Data Availability

All data produced in the present study are available upon reasonable request to the authors

## Author contributions

Brian C. Clark conceived the study, secured funding, and oversaw project administration. Brian C. Clark, Todd M. Manini, and Stuart J. Warden served as site principal investigators. Leatha A. Clark developed the study protocols and directed the central coordinating center. Janet E. Simon oversaw the statistical analyses. All authors contributed to study methodology and data interpretation. Brian C. Clark drafted the manuscript. All authors critically revised the manuscript and approved the final version for submission.

## Additional Information

This work was supported in part by a grant from the NIH’s National Institute on Aging (NIAR44AG058312). Study activities at the Indiana University site were partially supported by a grant from the National Institute of Arthritis and Musculoskeletal and Skin Diseases (NIAMS P30 AR072581).

## Acknowledgements

The authors thank Andrew Dick, an employee of OsteoDx, for his contributions to the development and technical refinement of the Cortical Bone Mechanics Technology (CBMT) platform, as well as for providing on-site support including device installation, operator training, and implementation across study sites. We also thank Max Stoeckel (OsteoDx), Massimo Ruzzene (Department of Mechanical Engineering, University of Colorado Boulder and consultant to OsteoDx), and Tony von Sadovszky (technical advisor to OsteoDx) for their contributions to the engineering development of the CBMT platform.

---

**Supplemental Table 1.** Inclusion and exclusion criteria for study participants in both target population groups (i.e., fragility fracture cases and controls).

| Inclusion Criteria Common Across Both Cases & Controls | Inclusion Criteria Specific to Fragility Fracture Cases | Inclusion Criteria Specific to Controls |
| --- | --- | --- |
| Females | Had experienced a fragility fracture of the arms (including wrist/carpal fractures) or legs (including hip, pelvis, and ankle fractures) after the age of 50 years. Insufficiency and stress fractures, as well as fractures of the spine, digits, toes, or face, were not included. A fragility fracture was operationally defined based on self-report of an arm or leg fracture caused by falls from a height <6 inches. A fragility fracture was not counted if it was associated with 1) running, bicycling or other similar fast-moving activity such as sports, 2) being struck by a falling or otherwise quickly moving heavy object, or 3) a motor vehicle accident. | Self-reported no history of fragility fractures at any time, or traumatic fractures at any site after age 40, excluding fractures of the digits, toes, or face. |
| 50-80 years of age |  | Does not self-report losing more than 3.8 cm (1.5 inches) in stature in the prior 15 years. |
| Self-reported that their last menses occurred at least 24-months prior to enrollment. |  |  |
| BMI between 18.5 and 35 kg/m<sup>2</sup>. |  |  |
| In the opinion of the site principal investigator the study participant is physically able to safely participate in the study activities |  |  |
| Able to provide informed consent |  |  |
| Exclusion Criteria Common Across Both Cases & Controls |  |  |
| Use of systemic glucocorticoids for more than 6-months in the prior one year |  |  |
| Self-reported diseases that could interfere with bone metabolism. For example, osteomalacia, bone cancer, myeloma, Paget's disease, hyper parathyroidism, hyperthyroidism not treated, severe renal (e.g., stage 4+ chronic kidney disease, history of dialysis, kidney transplant, etc.), hepatic insufficiency, or prolonged immobilization (more than 2 months in the previous year) |  |  |
| Self-reported type 1 diabetes |  |  |
| Self-reported being told by a physician that they have a terminal illness |  |  |
| Had had bilateral hip replacements |  |  |
| The subject was excluded if they answered yes to the following question: Do you have an active rotator cuff tear, had shoulder surgery in the past 12-months, or experience severe shoulder, wrist, or elbow joint pain on a regular basis? |  |  |
| Persons living in a nursing home (those living in assisted living or independent housing were not excluded) |  |  |
| Unable to communicate because of severe hearing loss or speech disorder |  |  |
| If, in the opinion of a site principal investigator, a study participant was inappropriate for the scientific purposes of this study. For instance, a high fall risk patient due to an existing neurological disorder (e.g., Parkinson's disease, ALS, etc.) would be excluded |  |  |
| Failure to provide informed consent |  |  |

**Supplemental Table 2.** Fracture Characteristics Among Case Participants (mean time since fracture: 5.0 ± 4.2 years).

| Fracture Location | Fracture Site | n (% of participants) |
| --- | --- | --- |
| **Upper Extremity** |  |  |
|  | Radius | 31 (28%) |
|  | Humerus | 19 (17%) |
|  | Ulna | 12 (11%) |
|  | Carpal | 11 (10%) |
|  | Clavicle | 1 (1%) |
| **Lower Extremity** |  |  |
|  | Fibula | 22 (20%) |
|  | Tibia | 12 (11%) |
|  | Hip (proximal femur) | 9 (8%) |
|  | Pelvis | 2 (2%) |
|  | Femur (distal) | 1 (1%) |
| **Total** |  |  |

*Percentages represent the proportion of case participants (n = 109) who experienced a fracture at each site. Because some individuals sustained more than one fracture, the total number of fractures exceeds the total number of cases and percentages sum to >100%.*

**Supplemental Table 3.** Osteoporosis pharmacotherapy exposure among study participants.

| **Drug Class** | **n (%)** |
| --- | --- |
| **Bisphosphonates** |  |
| Bisphosphonates — 1<sup>st</sup> Generation | 2 (1.4%) |
| Bisphosphonates — 2<sup>nd</sup> Generation | 76 (52.8%) |
| Bisphosphonates — 3<sup>rd</sup> Generation | 36 (25.0%) |
| **Monoclonal Antibodies** |  |
| Denosumab | 13 (9.0%) |
| Romosozumab | 5 (3.5%) |
| **PTH Analogs** | 10 (6.9%) |
| **SERM** | 1 (0.7%) |

*Percentages are expressed as proportion of total pharmacotherapy exposures.*

**Supplementary Table 4.**
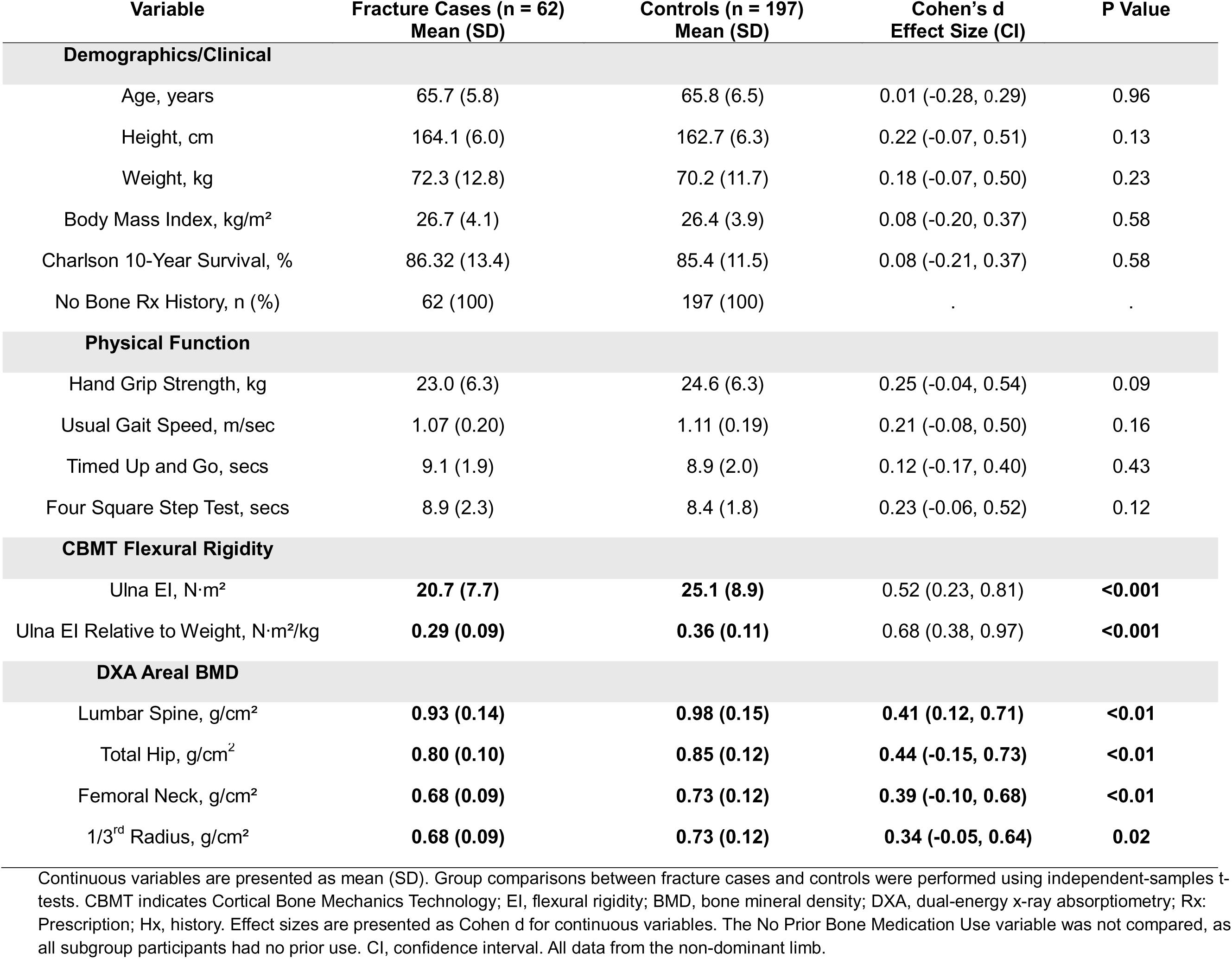
Participant Characteristics by Fracture Status in the Osteoporosis Treatment-Naïve Subset.

**Supplementary Table 5.** Participant Characteristics by Fracture Status in the Subset with Non-Osteoporotic BMD.

| Variable | Fracture Cases (n = 81)<br>Mean (SD) | Controls (n = 208)<br>Mean (SD) | Cohen's d<br>Effect Size (CI) | P Value |
| --- | --- | --- | --- | --- |
| **Demographics/Clinical** |  |  |  |  |
| Age, years | 67.0 (6.1) | 66.5 (6.4) | 0.09 (-0.16, 0.35) | 0.49 |
| Height, cm | 163.9 (6.1) | 163.1 (6.2) | 0.13 (-0.13, 0.38) | 0.33 |
| Weight, kg | 71.1 (12.9) | 70.9 (11.3) | 0.02 (-0.24, 0.28) | 0.89 |
| Body Mass Index, kg/m<sup>2</sup> | 26.4 (4.3) | 26.6 (3.8) | 0.05 (-0.21, 0.30) | 0.72 |
| Charlson 10-Year Survival, % | 83.7 (14.6) | 84.4 (11.5) | 0.06 (-0.20, 0.32) | 0.66 |
| No Bone Rx History, n (%) | **51 (63.0)** | **169 (81.3)** | **0.19** | **<0.01** |
| **Physical Function** |  |  |  |  |
| Hand Grip Strength, kg | **22.2 (6.7)** | **24.3 (6.5)** | **0.33 (0.07, 0.59)** | **0.02** |
| Usual Gait Speed, m/sec | 1.07 (0.20) | 1.11 (0.18) | 0.24 (-0.02, 0.49) | 0.07 |
| Timed Up and Go, secs | 9.2 (2.0) | 8.8 (1.7) | 0.20 (-0.06, 0.46) | 0.13 |
| Four Square Step Test, secs | **9.0 (2.2)** | **8.4 (1.7)** | **0.38 (0.12, 0.64)** | **0.004** |
| **CBMT Flexural Rigidity** |  |  |  |  |
| Ulna EI, N·m<sup>2</sup> | **20.6 (7.3)** | **25.6 (8.8)** | 0.60 (0.33, 0.87) | **<0.001** |
| Ulna EI Relative to Weight, N·m<sup>2</sup>/kg | **0.29 (0.10)** | **0.36 (0.11)** | 0.65 (0.38, 0.92) | **<0.001** |
| **DXA Areal BMD** |  |  |  |  |
| Lumbar Spine, g/cm<sup>2</sup> | **0.97 (0.12)** | **1.01 (0.14)** | **0.28 (-0.02, 0.55)** | **0.03** |
| Total Hip, g/cm<sup>2</sup> | **0.82 (0.09)** | **0.86 (0.09)** | **0.41 (-0.15, 0.68)** | **0.02** |
| Femoral Neck, g/cm<sup>2</sup> | **0.70 (0.08)** | **0.74 (0.10)** | **0.43(-0.17, 0.70)** | **0.02** |
| 1/3<sup>rd</sup> Radius, g/cm<sup>2</sup> | 0.63 (0.10) | 0.65 (0.08) | 0.23 (-0.03 0.49) | 0.08 |

Continuous variables are presented as mean (SD). Group comparisons between fracture cases and controls were performed using independent-samples t-tests. CBMT indicates Cortical Bone Mechanics Technology; EI, flexural rigidity; BMD, bone mineral density; DXA, dual-energy x-ray absorptiometry; Rx: Prescription; Hx, history. Effect sizes are presented as Cohen d for continuous variables and as the Phi coefficient for the No Prior Bone Medication Use variable. CI, confidence interval. All data from the non-dominant limb.

**Supplemental Table 6.** Univariable Logistic Regression and AUC Results for Age and BMI in Discriminating Fracture Status in the Full Sample. Threshold-based metrics (cutoff, sensitivity, specificity, PPV, NPV) are omitted for predictors with non-significant discriminatory performance (AUC P > .05).

| Logistic Regression Results |  |  |  | ROC Curve Results |  |  |  |  |  |  |  |
| --- | --- | --- | --- | --- | --- | --- | --- | --- | --- | --- | --- |
| Predictor | OR<br>Per SD | 95% CI | P-Value | AUC | 95% CI | P-Value | Cutoff | Sensitivity<br>(%) | Specificity<br>(%) | PPV<br>(%) | NPV<br>(%) |
| Age | 1.09 | 0.85, 1.42 | 0.51 | 0.53 | 0.42, 0.65 | 0.58 | – | – | – | – | – |
| BMI | 0.93 | 0.93, 0.94 | 0.62 | 0.48 | 0.35, 0.61 | 0.67 | – | – | – | – | – |

Odds ratios (ORs) are expressed per 1–standard deviation (SD) decrease in the predictor. ROC = Receiver operating characteristic curve. AUC = area under the ROC curve.

**Supplemental Table 7.** Multivariable Logistic Regression Models Predicting Fracture Status Using CBMT-Derived Flexural Rigidity Normalized to Body Weight with Adjustment for Age, BMI, and DXA-Derived BMD in the Full Sample.

|  |  | Logistic Regression Results |  |  | ROC Curve Results |  |  |
| --- | --- | --- | --- | --- | --- | --- | --- |
| Model | Predictors | OR Per SD | 95% CI | P-Value | AUC | 95% CI | P-Value |
| Relative CBMT EI Only (Univariable) |  |  |  |  | 0.80 | 0.70–0.89 | < 0.001 |
|  | Relative CBMT EI | 0.57 | 0.42, 0.80 | < 0.001 |  |  |  |
| Relative CBMT EI + Age and BMI |  |  |  |  | 0.77 | 0.67-0.87 | <0.001 |
|  | Relative CBMT EI | 0.56 | 0.39, 0.81 | <0.001 |  |  |  |
|  | Age | 1.06 | 0.79, 1.42 | 0.69 |  |  |  |
|  | BMI | 0.81 | 0.60, 1.10 | 0.17 |  |  |  |
| Relative CBMT EI + Lumbar Areal BMD |  |  |  |  | 0.78 | 0.68, 0.87 | < 0.001 |
|  | Relative CBMT EI | 0.57 | 0.40, 0.82 | < 0.001 |  |  |  |
|  | Lumbar Areal BMD | 0.83 | 0.62, 1.11 | 0.20 |  |  |  |
| Relative CBMT EI + Total Hip Areal BMD |  |  |  |  | 0.78 | 0.69, 0.88 | < 0.001 |
|  | Relative CBMT EI | 0.60 | 0.42, 0.85 | < 0.01 |  |  |  |
|  | Total Hip Areal BMD | 0.71 | 0.53, 0.96 | 0.02 |  |  |  |
| Relative CBMT EI + Femoral Neck Areal BMD |  |  |  |  | 0.76 | 0.66, 0.87 | < 0.001 |
|  | Absolute CBMT EI | 0.61 | 0.42, 0.87 | < 0.01 |  |  |  |
|  | Femoral Neck Areal BMD | 0.73 | 0.53, 0.98 | 0.05 |  |  |  |
| Relative CBMT EI + 1/3rd Radius Areal BMD |  |  |  |  | 0.78 | 0.68, 0.87 | < 0.001 |
|  | Absolute CBMT EI | 0.60 | 0.42, 0.86 | < 0.01 |  |  |  |
|  | 1/3rd Radius Areal BMD | 0.80 | 0.58, 1.12 | 0.19 |  |  |  |

Odds ratios (ORs) are expressed per 1–standard deviation (SD) decrease in the predictor. ROC = Receiver operating characteristic curve. AUC = area under the ROC curve. CBMT = Cortical Bone Mechanics Technology; EI = Flexural rigidity; BMD = bone mineral density. All data from the non-dominant limb.

